# Blood biomarker changes in response to low-dose oral ketamine treatment in adults with major depressive disorder (MDD) and post-traumatic stress disorder (PTSD)

**DOI:** 10.64898/2026.08.12.26360216

**Authors:** Annette M. Braxton, Christina Driver, Daniel F. Hermens, Bonnie L. Quigley

**Affiliations:** National PTSD Research Centre at the Thompson Institute, University of the Sunshine Coast, Birtinya, QLD, 4575, Australia; Centre for Bioinnovation, University of the Sunshine Coast, Sippy Downs, QLD, 4556, Australia

**Keywords:** Depression, major depressive disorder, PTSD, post-traumatic stress disorder, ketamine, biomarker

## Abstract

Major depressive disorder (MDD) and post-traumatic stress disorder (PTSD) are prevalent, chronic, and disabling mental health conditions which are difficult to treat. Ketamine has demonstrated effect for improving depression and PTSD symptoms independently but reports often overlook focused comorbid improvement of these symptoms within individuals. To address this, we assessed blood-based biomarker and psychological changes following low-dose oral ketamine treatment for adults with MDD alone (n=14) and comorbid MDD+PTSD (n=21). Before treatment, the MDD clinical group presented with more severe depression, lower serotonin levels and higher kynurenine levels than the MDD+PTSD group. Post-treatment there were no detectable differences in the biological response between clinical groups, with combined analysis revealing common decreases in circulating brain-derived neurotrophic factor (BDNF) and vascular endothelial growth factor (VEGF-A). Additionally, post-treatment, both clinical groups showed improvements in anxiety, stress, social functioning, suicidal ideation, and general well-being measures, as well as individual improvements in depression scores and PTSD symptoms in the MDD- and PTSD-containing groups, respectively. Collectively, this study presents additional evidence that low-dose oral ketamine treatment can be effective for MDD and MDD+PTSD, individually and comorbidly, and that both MDD and PTSD clinical groups responded in the same biological manner.

**Highlights:**

- Low dose oral ketamine can be effective in treating MDD and PTSD concurrently
- Blood biomarkers BDNF and VEGF decreased with symptom improvement
- No detectable difference in oral ketamine treatment response by clinical diagnosis

## 1. Introduction

Major depressive disorder (MDD) and post-traumatic stress disorder (PTSD) are prevalent, chronic, debilitating, and disabling affective and trauma/stressor-related disorders, respectively, which are difficult to treat and have high rates of morbidity and mortality. Diagnosis of these disorders contributes to worldwide disease and economic burden, negatively affecting personal relationships, employment, psychosocial functioning, and quality of life (Koenen et al., 2017; Lu et al., 2024). Despite psychotherapeutic and pharmacotherapeutic interventions, failure to respond to therapy occurs in 30-50% of people with MDD or PTSD (Semmlinger et al., 2024; Sicras-Mainar et al., 2012). Selective serotonin reuptake inhibitors (SSRIs) and serotonin-norepinephrine reuptake inhibitors (SNRIs) are first-line treatments for MDD, and SSRIs are the only United States (US) Food and Drug Administration (FDA) approved pharmacotherapeutics for PTSD. SSRIs and SNRIs have shown only moderate efficacy in both MDD and PTSD, may have delayed therapeutic actions often resulting in early discontinuation, and adverse side-effects with long-term use (Nierenberg et al., 2008; Van Etten and Taylor, 1998). This has created a critical need for urgent clinical advancements of novel, rapid, robust, and efficacious pharmacotherapeutic treatments with transdiagnostic potential.

Ketamine is a non-competitive *N*-methyl-d-aspartate (NMDA) receptor antagonist, which began showing promise as a rapid and robust antidepressant in people with MDD twenty-six years ago (Berman et al., 2000; Driver et al., 2022). Since then, multiple controlled trials have demonstrated ketamine’s rapid antidepressant response in individuals with MDD, treatment-resistant depression, and suicidal ideation (Glue et al., 2024; Kryst et al., 2020; Parikh et al., 2024; Seraj et al., 2025). In parallel, evidence has accumulated that similar positive treatment outcomes can be achieved with ketamine for the treatment of other disorders, such as PTSD (Ragnhildstveit et al., 2023). Ketamine is highly liposoluble and multiple routes of administration may be used, with various bioavailability ranges: intravenous (IV) (100%), intramuscular (93%), subcutaneous (90-95%), intranasal (45-50%), sublingual (20-30%), and oral (17-29%) (Gibula-Tarlowska et al., 2025; McIntyre et al., 2021). IV ketamine is currently the most widely studied and utilised, providing a rapid, but often short-lived therapeutic effect and higher peak concentrations, leading to acute changes in biomarkers (Gibula-Tarlowska et al., 2025). However, its cost and hospital setting delivery can limit patient and clinician accessibility in real-world settings. In comparison, oral ketamine can easily be delivered in a range of settings, is non-invasive, cost effective, and may increase patient and clinician accessibility. Oral ketamine administration has low ketamine bioavailability due to first-pass hepatic metabolism, which results in delayed onset of therapeutic effects and lower circulating concentrations, leading to slower changes in biomarkers (Dutton et al., 2023; Peltoniemi et al., 2016). Importantly, norketamine blood concentrations (a primary metabolite of ketamine), have been shown to be higher following oral administration (Dutton et al., 2023), resulting in a longer elimination time than ketamine, producing sustained therapeutic effects (Otto et al., 2026; Zanos et al., 2018). Taken together, oral ketamine provides a viable, potentially effective therapy for MDD and PTSD, however, there is still a research gap into oral ketamine outcomes (both biological and clinical) for comorbid MDD+PTSD.

In addition to assessing clinical outcomes from ketamine treatment, biomarkers may play a key role in understanding whether individuals with MDD and/or PTSD will improve with ketamine therapy (Meshkat et al., 2023), due to underlying neurobiological mechanisms and changes that are common among these disorders. Beyond the traditional immune markers, putative ketamine-related biomarkers have included the neurotrophic factors, brain-derived neurotrophic factor (BDNF) and vascular endothelial growth factor A (VEGF), as well as the neurotransmitter/hormone, serotonin. BDNF plays a vital role in neurogenesis, neuroplasticity and neuroprotection, which is essential for learning and memory (Bathina and Das, 2015), while VEGF supports vasculogenesis, angiogenesis, neurogenesis, and neuroprotection (Wiszniak and Schwarz, 2021). It has been suggested that BDNF and VEGF work together to promote neuroplasticity and neuroprotection and that reduced or abnormal levels of either one or both of these neurotrophic factors may be involved in the development and maintenance of psychiatric disorders (Matsuzaka and Yashiro, 2025). Recent meta-analysis of BDNF levels from multiple psychiatric conditions suggest that BDNF levels appear reduced in MDD but elevated in PTSD (Zou et al., 2024), while other reviews/studies report elevated VEGF levels in people with MDD or PTSD compared with healthy controls (Matsuzaka and Yashiro, 2025; Quigley et al., 2025b). Alternatively, serotonin is an important neurotransmitter and peripheral hormone which is responsible for the regulation of stress responses, mood, behaviour, sleep, memory, and appetite (Kanova and Kohout, 2021). Serotonin is synthesised from tryptophan, with tryptophan metabolism believed to be involved in the pathophysiology of neuropsychiatric disorders due to the synthesis of both serotonin and kynurenine, with the balance between these two pathways playing an important role in physiological homeostasis (Correia and Vale, 2022). Chronic stress responses are indicated in MDD and PTSD and may shift the balance of the serotonin and kynurenine pathways, which can influence the development of MDD (Correia and Vale, 2022). While there have been limited investigations into these biomarkers related to MDD or PTSD separately, there is currently a lack of data informing on how these important biomarkers respond to oral ketamine treatment in the presence of comorbid MDD and PTSD.

Therefore, the aim of this study was to evaluate biological and psychological outcomes in response to low-dose ketamine treatment among participants diagnosed with MDD (without PTSD) and participants diagnosed with comorbid MDD+PTSD. These analyses assessed the acute (one-week post-treatment) biological and psychological responses to oral ketamine therapy overall and by MDD/PTSD status, as per a trial protocol (Quigley et al., 2025a). The goal was to determine whether there were significant biological and psychological differences between the two disorder groups, with various MDD/PTSD clinical profiles, before treatment and if there were characteristic differences between clinical groups after oral ketamine therapy.

## 2. Materials and methods

### 2.1 Study participants

The present study is a subset analysis of participants enrolled in clinical trials at the Thompson Institute, University of the Sunshine Coast (UniSc), Queensland, Australia (Clinical Trials Registry of Australia and New Zealand ACTRN12621000429853 and ACTRN12618001965291; (Quigley et al., 2025a)). All participants included in the study had received a confirmed diagnosis of MDD (n=14) or MDD+PTSD (n=21), from a qualified psychiatrist. This study examined the use of oral ketamine as an augmentation treatment for MDD and MDD+PTSD. All experimental protocols were approved by the Metro North Health Human Ethics Committee (HREC/2019/QPCH/53437 and HREC/18/QPCH/288) and the UniSC Ethics Committee (A211537 and A181190). All procedures adhered strictly to pertinent guidelines and regulations, and written informed consent was obtained from every participant (Quigley et al., 2025a). (Participant recruitment, inclusion and exclusion criteria are shown in **Supplementary Table 1**).

Recruited participants included in the studies were aged 18 years or older, consisting of males (MDD, n=6 and PTSD, n=7) and females (MDD n=8 and PTSD n=14). Diagnosis of MDD and/or PTSD were obtained by utilising the Hamilton Depression Rating Scale (HAM-D) (Hamilton, 1960) for depression or the Montgomery-Asberg Depression Rating Scale (MADRS) (Montgomery and Asberg, 1979), along with the Clinician Administered PTSD Scale for DSM-5 (CAPS-5) (Weathers et al., 2018) for PTSD. To maximise sample number for analysis, participants were included in the study if baseline data was available in addition to follow-up 1 (FUP1) (one-week post-treatment). This created a combined study sample set of MDD participants with n=14 at baseline, n=11 at FUP1, and MDD+PTSD participants with n=21 at baseline, n=20 at FUP1.

### 2.2 Ketamine trial procedures

Participants physical health was evaluated with a physical examination, pathology blood tests, and reviews of individual medical history. Throughout the treatment and follow-up phase, participants continued taking regularly prescribed medications as directed by individual treating physicians. Thus, most participants within these studies were taking concurrent pharmacotherapeutics, such as SSRIs, SNRIs, mood stabilisers or antipsychotics. Demographic data, concurrent pharmacotherapeutics, and comorbid medical conditions are shown in **Supplementary Table 2** (**A and B**).

The studies were conducted at the Thompson Institute, University of the Sunshine Coast, from February 2021-December 2024. A flexible course of low dose oral (racemic) ketamine (titrated from 0.5 mg/kg to a maximum of 3.0 mg/kg) was administered once-weekly for six weeks, followed by a 1-week follow-up (FUP1) without ketamine (Quigley et al., 2025a).

### 2.3 Blood collection

A phlebotomist collected non-fasting whole blood samples at all timepoints. Serum-separator tubes (SST) were used for serum. Collection tubes remained at room temperature for a minimum of 30 minutes to allow for clotting. Tubes were then centrifuged at 2465 x g for 15 minutes at 4°C. Samples were all processed within four hours of collection, with serum separated, aliquoted, then stored at −80°C.

### 2.4 Biomarker quantification

Serum samples were thawed on ice and centrifuged at 10,000 x g for 10 minutes at 4°C to remove precipitates before analysis. Peripheral biomarkers were quantified; BDNF (ProcartaPlex simplex assay, ThermoFisher Scientific, Australia; lower limit of quantification (LLOQ) of 2.03 pg/ml), and VEGF-A (ProcartaPlex simplex assay, ThermoFisher Scientific; LLOQ of 4.88 pg/ml) by xMAP microsphere-based assay (Luminex, ThermoFisher Scientific).

Tryptophan and kynurenine (Immusmol, France; LLOQ of 2.5 µg/ml and LLOQ of 100 ng/ml respectively) and serotonin (Fast Track kit, Immusmol, France; LLOQ of 15 ng/ml), were measured utilizing enzyme-linked immunosorbent assay (ELISA, Immusmol or ThermoFisher Scientific).

### 2.5 Clinical (psychological) scales data collection

The following clinical scales were assessed at baseline and the follow-up timepoint: Depression, Anxiety, and Stress Scale (DASS-21) (Lovibond and Lovibond, 1995), a 21-item-self-report measure to identify depression, anxiety, and stress symptomatology; Social and Occupational Functioning Assessment Scale (SOFAS) (Goldman et al., 1992), a clinician-rated single-item scale used to indicate an individuals’ level of social and occupational functioning; Beck Scale for Suicidal Ideation (BSS) (Beck et al., 1979), a 21-item clinician-rated scale examining suicidal intent; and the World Health Organisation Well-Being Index (WHO-5) (Bech et al., 2003), a 5-item self-report to assess mental well-being. The primary outcome measure for the MDD group utilised HAM-D (Hamilton, 1960), a 17-item clinician-rated scale used to measure the severity of depressive symptoms, whilst the PTSD group utilised the Montgomery-Asberg Depression Rating Scale (MADRS) (Montgomery and Asberg, 1979), a 10-item clinician-rated scale used to measure the severity of depressed symptomology; and the Post-traumatic Stress Disorder Checklist for DSM-5 (PCL-5) (Blevins et al., 2015), a 20-item self-report assessment used to measure severity of PTSD symptoms.

### 2.6 Statistical analysis

R Statistical Software (v4.3.3) was used for statistical analyses (R Core Team, 2024). The study used two independent groups (MDD and MDD+PTSD), often with non-normal distributions as determined by Shapiro-Wilk test (Shapiro and Wilk, 1965), and had small unequal sample sizes. As such, the Welch two-sample t-test (Welch, 1947) or the Whitney U/Wilcoxon test (Mann and Whitney, 1947) was used as appropriate to determine overall differences between groups and to make multiple pairwise comparisons, with corrected p-values. The combination of the Welch two-sample t-test and Whitney U/Wilcoxon test with continuity correction was used to compare differences in baseline and the follow-up timepoint for all study parameters (BDNF, VEGF-A, tryptophan, serotonin, and kynurenine; DASS-21-subscales of depression, anxiety and stress, SOFAS, BSS, WHO-5, PCL-5, MADRS, and HAM-D). Comparison of study groups by sex was completed using the Fisher Exact test (Fisher, 1934), while comparisons of study groups by age was completed using the student’s t-test (Student, 1908).

Changes in study parameters over the course of the ketamine trial were assessed by linear mixed model (LMM) analysis using lme4 (Bates et al., 2015) and lmerTest (Kuznetsova et al., 2017) and visualised using ggplot2 (Wickham, 2016). Significance thresholds were set at p ≤ 0.05.

## 3. Results

A total of 35 participants were analysed (**Table 1**). The mean age in the study was 39.75 ± 14.15 years. MDD participants were found to be significantly younger than MDD+PTSD participants (**Table 1**). The total sample included 13 males (37%) and 22 females (63%), a gender distribution that did not significantly differ between the two groups.

**Table 1:** Demographics at baseline Table 1: Mean demographics, biological levels and psychological scores (± standard deviation) of the two clinical groups (MDD only and MDD+PTSD) at baseline. Significant differences are highlighted in bold. *Welch two-sample t-test or Mann Whitney U/Wilcoxon rank sum test with continuity correction used, as appropriate. nd = not done.

| Characteristic | MDD | MDD+PTSD | Test statistic* | P-value |
| --- | --- | --- | --- | --- |
| Number of participants | 14 | 21 | n/a | n/a |
| Male/Female (%M) | 6M/8F (43%) | 7M/14F (33%) | n/a | 0.712 |
| Age (years) | $32.8 \pm 13.6$ | $46.7 \pm 11.8$ | $t(25.3) = -3.12$ | **0.004** |
| BDNF (pg/ml) | $129.75 \pm 101.60$ | $230.21 \pm 340.76$ | W = 134 | 0.678 |
| VEGF-A (pg/ml) | $279.56 \pm 181.69$ | $298.30 \pm 168.85$ | W = 134 | 0.678 |
| Tryptophan (μg/ml) | $12.70 \pm 3.71$ | $10.67 \pm 3.63$ | W = 195 | 0.110 |
| Serotonin (ng/ml) | $37.50 \pm 39.48$ | $88.60 \pm 76.18$ | W = 73 | **0.013** |
| Kynurenine (ng/ml) | $732.95 \pm 288.98$ | $517.42 \pm 296.84$ | $t(28.5) = 2.14$ | **0.041** |
| DASS-21 depression | $28.71 \pm 8.43$ | $19.81 \pm 9.63$ | $t(30.5) = 2.89$ | **0.007** |
| DASS-21 anxiety | $16.57 \pm 8.79$ | $14.57 \pm 8.74$ | $t(27.9) = 0.66$ | 0.514 |
| DASS-21 stress | $21.14 \pm 7.95$ | $19.90 \pm 8.95$ | $t(30.2) = 0.43$ | 0.671 |
| SOFAS | $55.00 \pm 10.92$ | $59.05 \pm 17.29$ | W = 124.5 | 0.442 |
| BSS | $11.75 \pm 9.46$ | $5.29 \pm 15.77$ | W = 174.5 | 0.072 |
| WHO-5 | $18.57 \pm 15.58$ | $27.81 \pm 15.77$ | W = 92 | 0.064 |
| PCL-5 | nd | $40.95 \pm 11.77$ | nd | nd |
| MADRS | nd | $33.43 \pm 7.55$ | nd | nd |
| HAM-D | $29.43 \pm 5.79$ | nd | nd | nd |

### 3.1 Baseline comparison of biological markers for study groups

Baseline biological markers showed a significant difference in both serotonin and kynurenine levels between the MDD and MDD+PTSD groups, with lower serotonin and higher kynurenine levels detected in the MDD group (**Table 1**). No significant differences between the clinical groups for BDNF, VEGF-A or tryptophan levels were detected at baseline (**Table 1**).

### 3.2 Baseline comparison of psychological scales for study groups

Baseline psychological scales showed a significant difference between the MDD group and the MDD+PTSD group for the DASS-21-depression subscale, with more severe depressive symptoms in the MDD group (**Table 1)**. No other significant differences were observed between the clinical groups for other psychological scales (**Table 1**).

### 3.3 Biological markers following ketamine treatment by study group

The biological markers tested (BDNF, VEGF-A, tryptophan, serotonin, and kynurenine) did not show any group-level significant differences/changes, regardless of clinical status, following treatment with oral ketamine. (**Table 2A and B**).

**Table 2:** Change in parameters by study group (A) MDD and (B) MDD+PTSD A. Table 2: Mean demographics, biological levels and psychological scores (± standard deviation) of the two clinical groups (MDD only and MDD+PTSD) at baseline and FUP1. Significant differences are highlighted in bold. *Welch two-sample t-test or Mann Whitney U/Wilcoxon rank sum test with continuity correction used, as appropriate.

| Characteristic | MDD Baseline | MDD FUP1 | Test statistic* | P-value |
| --- | --- | --- | --- | --- |
| BDNF (pg/ml) | 129.75 ± 101.60 | 69.67 ± 43.88 | W = 99 | 0.244 |
| VEGF-A (pg/ml) | 279.56 ± 181.69 | 252.03 ± 269.22 | W = 97 | 0.292 |
| Tryptophan (µg/ml) | 12.70 ± 3.71 | 12.43 ± 3.37 | t(22.5) = 0.19 | 0.851 |
| Serotonin (ng/ml) | 37.50 ± 39.48 | 26.91 ± 47.01 | W = 94.5 | 0.349 |
| Kynurenine (ng/ml) | 732.95 ± 288.98 | 567.79 ± 213.54 | t(22.9) = 1.64 | 0.114 |
| DASS-21 depression | 28.71 ± 8.43 | 13.64 ± 14.31 | t(15.3) = 3.10 | **0.007** |
| DASS-21 anxiety | 16.57 ± 8.79 | 5.27 ± 6.59 | W = 135.5 | **0.001** |
| DASS-21 stress | 21.14 ± 7.95 | 8.73 ± 5.68 | t(22.8) = 4.55 | **<0.001** |
| SOFAS | 55.00 ± 10.92 | 82.73 ± 17.37 | W = 17 | **0.001** |
| BSS | 11.75 ± 9.46 | 8.27 ± 8.51 | W = 86 | 0.227 |
| WHO-5 | 18.57 ± 15.58 | 45.82 ± 27.73 | W = 28 | **0.008** |
| HAM-D | 29.43 ± 5.79 | 7.36 ± 5.80 | W = 154 | **<0.001** |

**Table 2:** Mean demographics, biological levels and psychological scores (± standard deviation) of the two clinical groups (MDD only and MDD+PTSD) at baseline and FUP1.
| Characteristic | MDD+PTSD Baseline | MDD+PTSD FUP1 | Test statistic* | P-value |
| --- | --- | --- | --- | --- |
| BDNF (pg/ml) | 230.21 ± 340.76 | 187.81 ± 240.51 | W = 238 | 0.447 |
| VEGF-A (pg/ml) | 298.30 ± 168.85 | 278.30 ± 156.79 | t(39.0) = 0.39 | 0.696 |
| Tryptophan (µg/ml) | 10.67 ± 3.63 | 11.22 ± 3.88 | t(38.5) = -0.47 | 0.638 |
| Serotonin (ng/ml) | 88.60 ± 76.18 | 84.09 ± 86.64 | W = 227.5 | 0.658 |
| Kynurenine (ng/ml) | 517.42 ± 296.84 | 531.48 ± 256.91 | W = 201 | 0.825 |
| DASS-21 depression | 19.81 ± 9.63 | 10.42 ± 9.54 | W = 310.5 | **0.003** |
| DASS-21 anxiety | 14.57 ± 8.74 | 6.11 ± 8.83 | W = 318 | **0.001** |
| DASS-21 stress | 19.90 ± 8.95 | 11.58 ± 9.13 | t(37.4) = 2.91 | **0.006** |
| SOFAS | 59.05 ± 17.29 | 73.16 ± 16.00 | t(38.0) = -2.68 | **0.011** |
| BSS | 5.29 ± 4.62 | 2.74 ± 4.41 | W = 277.5 | **0.032** |
| WHO-5 | 27.81 ± 15.77 | 44.84 ± 19.49 | W = 97 | **0.006** |
| PCL-5 | 40.95 ± 11.77 | 20.58 ± 15.90 | t(33.0) = 4.57 | **<0.001** |
| MADRS | 33.43 ± 7.55 | 11.72 ± 12.07 | W = 346 | **<0.001** |

### 3.4 Psychological scales following ketamine treatment by study group

Following oral ketamine administration, most psychological scales showed improvements across clinical groups, regardless of group status. Significant improvements were shown by the MDD and MDD+PTSD groups from baseline-FUP1 in all subscales of DASS-21 (depression, anxiety and stress), WHO-5 and SOFAS scores. Significant improvements were shown in the MDD+PTSD group from baseline-FUP1 in PCL-5 and MADRS scores, and the MDD group showed significant improvements in HAM-D scores from baseline-FUP1. The only scale that did not show significant improvement for the MDD group was the BSS score. However, significant improvement was shown in the MDD+PTSD group’s BSS score (**Table 2 A and B**).

### 3.5 Changes in study parameters with participants as a combined group

LMMs for the MDD and MDD+PTSD clinical groups were used to investigate the change within individuals for each study parameter. Accounting for participant age, models determined that neither clinical group nor sex were significant factors accounting for changes in the study parameters. Given this lack of unique treatment outcome based on MDD or PTSD status (or sex), all participants were further analysed as a single oral ketamine treatment group. **Table 3** reports fixed effect coefficient (β) changes from baseline-FUP1 for each study parameter. Positive coefficients indicate increases from baseline in that parameter while negative coefficients indicate decreases from baseline. The combined oral ketamine treatment group analysis showed significant change differences were observed by LMM analysis for the biological markers BDNF and VEGF-A (**Table 3**). BDNF and VEGF-A within this combined cohort had significantly reduced levels from baseline-FUP1. Additionally, significant improvements were shown in all psychological scales following oral ketamine treatment regardless of MDD or PTSD diagnosis (**Table 3**).

**Table 3:** Linear Mixed Model changes by group for baseline – FUP1. Table 3: β = Fixed effect coefficients (represents the change in the indicated parameter in that timeframe; positive numbers indicate increases, negative numbers indicate decreases), p = p-value. Significant changes from baseline are highlighted in bold. nd = not done.

| Parameters | MDD |  | MDD+PTSD |  | All oral ketamine treated |  |
| --- | --- | --- | --- | --- | --- | --- |
| | $\beta$ | p-value | $\beta$ | p-value | $\beta$ | p-value |
| BDNF (pg/ml) | -48.3 | 0.066 | -50.7 | 0.081 | **-50.5** | **0.013** |
| VEGF-A (pg/ml) | -39.7 | 0.114 | -19.4 | 0.177 | **-27.1** | **0.033** |
| Tryptophan (μg/ml) | -0.2 | 0.867 | 0.5 | 0.536 | 0.3 | 0.667 |
| Serotonin (ng/ml) | 0.9 | 0.896 | 4.8 | 0.548 | 3.3 | 0.542 |
| Kynurenine (ng/ml) | -168.3 | 0.077 | 4.1 | 0.950 | -57.5 | 0.297 |
| DASS-21 depression | **-14.3** | **$1.92 \times 10^{-4}$** | **-10.3** | **$1.31 \times 10^{-4}$** | **-11.8** | **$2.47 \times 10^{-8}$** |
| DASS-21 anxiety | **-12.2** | **$1.93 \times 10^{-4}$** | **-7.9** | **0.002** | **-9.6** | **$6.29 \times 10^{-7}$** |
| DASS-21 stress | **-12.5** | **$3.52 \times 10^{-5}$** | **-8.7** | **$9.13 \times 10^{-4}$** | **-10.1** | **$1.49 \times 10^{-7}$** |
| SOFAS | **28.0** | **$2.94 \times 10^{-5}$** | **15.8** | **0.003** | **20.3** | **$2.47 \times 10^{-6}$** |
| BSS | **-5.7** | **0.013** | **-2.6** | **0.041** | **-3.7** | **0.001** |
| WHO-5 | **25.8** | **$3.97 \times 10^{-5}$** | **19.2** | **$1.13 \times 10^{-4}$** | **21.7** | **$1.81 \times 10^{-8}$** |
| PCL-5 | nd | nd | **-21.7** | **$1.63 \times 10^{-6}$** | nd | nd |
| MADRS | nd | nd | **-22.3** | **$7.79 \times 10^{-9}$** | nd | nd |
| HAM-D | **-22.1** | **$4.59 \times 10^{-11}$** | nd | nd | nd | nd |

## 4. Discussion

The current study has contributed to the growing evidence base for the use of low-dose oral ketamine as a viable treatment for individuals with a standalone diagnosis of MDD, and comorbid MDD+PTSD, and supports its efficacy. Additionally, analysis showed that oral ketamine appeared to have the same biological effect on blood biomarkers in both MDD and MDD+PTSD participants (i.e., regardless of clinical diagnosis). Our integrated analysis found baseline differences in serotonin and kynurenine biomarkers between clinical groups, and combined treatment analysis revealed shared reduced BDNF and VEGF-A levels across both groups. As well, robust improvements in both clinician-administered and self-reported psychological measures were reported, with analyses showing significant improvements in severity of symptoms, overall functioning, and psychological wellbeing. These findings align with previous studies on oral ketamine administration which show significant improvements in depressive symptoms in MDD (Glue et al., 2020; Glue et al., 2024; Kumar et al., 2024; Veraart et al., 2023) and PTSD symptoms in PTSD (Quigley et al., 2025a). A novel finding that this specific study adds is detailed characterisation of several blood biomarkers showing similar biological response to oral ketamine in the context of these two distinct mental health diagnoses.

The baseline data analysis for this study cohort detected a significant difference in age between the two clinical groups in this study, with MDD participants being younger. Interestingly, Australian statistics currently suggest that both MDD and PTSD have higher rates in younger adults compared to older adults (Australian Bureau of Statistics 2020-21; Phoenix Australia, 2020). The older age of participants in these studies with PTSD may have reflected a differing prevalence and/or need in the local community (southeast Queensland, Australia), where recruitment was open to the public and participants were self-selecting. This demographic difference would be interesting to pursue in future studies.

There were significant differences detected in serotonin and kynurenine levels at baseline between the MDD and the MDD+PTSD groups, with lower serotonin and higher kynurenine levels seen in the former. This reciprocal relationship was not surprising, given the shared tryptophan precursor pathway for serotonin and kynurenine, and that lower serotonin levels have been consistently associated with MDD (Colle et al., 2020; Correia and Vale, 2022).

The only significant difference within the psychological scale scores between clinical groups at baseline was the DASS-21 depression subscale, with participants from the MDD group reporting more severe depression symptoms compared to the MDD+PTSD group. This contradicts reports investigating the severity of depression alone or co-morbid with PTSD in military veterans, which found that MDD+PTSD patients reported more severe depression compared to MDD patients (Campbell et al., 2007). Whether the outcome reflects a difference inherent to our large civilian study cohort or simply a unique characteristic of this group of individuals remains to be further explored.

Biomarker changes following ketamine treatment at the combined sample level showed BDNF and VEGF-A levels significantly reduced post oral ketamine treatment. This outcome contrasts with IV ketamine studies that report increases, particularly in relation to BDNF levels during successful treatment of MDD (Haile et al., 2014; Kang and Vazquez, 2022; Zheng et al., 2021). Whether the difference in BDNF change in the MDD group is related to the route of ketamine administration (oral verses IV; leading to different primary and secondary active ketamine metabolites) is a question for future pharmacokinetic studies. Additionally, in clinical groups where MDD (characterised by lower BDNF levels) is comorbid with PTSD (characterised by higher BDNF levels) (Zou et al., 2024), it is still unclear whether increases or decreases in overall BDNF levels would be most beneficial. As such, we cannot definitively state the positive mental health outcomes detected in this study, which includes both MDD and PTSD, were related to the decreases in BDNF and/or VEGF-A levels or whether they were achieved despite these decreases in blood biomarkers. Investment into more targeted research of BDNF (and VEGF-A) and the potential mechanism of oral ketamine action is needed with larger samples for clarification.

Oral ketamine treatment showed that both clinical groups in this study had improvements in common psychological scales for stress, functioning and well-being. These findings are consistent with previous research with ketamine treatment for MDD or PTSD, where IV ketamine therapy often shows a significant acute response in efficacy (Feder et al., 2021; Kryst et al., 2020). Our results also align with the outcomes from (Johnson et al., 2025), which showed comorbid MDD+PTSD observed significant improvements in both depression and PTSD psychological scores following four IV ketamine infusions across 8-10 days. Further transdiagnostic potential for ketamine was reported by (Daly et al., 2021), which showed esketamine nasal spray treatment for MDD and comorbid MDD+anxiety showed significant reductions in MADRS scores, regardless of diagnosis. Taken together, previous studies and our current results support ketamine therapy through multiple delivery routes for symptom improvement transdiagnostically across multiple psychiatric disorders, including with comorbidity. The overlap in efficacy is most likely the result of similar symptomology and molecular pathways being shared across many neuropsychiatric disorders (Davidson et al., 2022).

There were limitations of this analysis that should be acknowledge. The small sample size, particularly for the MDD group, may have underpowered parts of the study by limiting the variability and representativeness of the cohort studied and impacted the statistical power. Additionally, different primary clinical depression scales were used for the MDD group (HAM-D) and the MDD+PTSD group (MADRS), which limited direct comparisons between groups. Moreover, both trials were open label with no placebo group. Finally, most study participants were on concurrent medications that may have affected the ketamine treatment-related outcomes. A review by (Veraart et al., 2021), showed that prescribed medications for depression are often combined with ketamine to prevent relapse, with most published evidence analysing IV administration with a range of antidepressant medications which suggest synergistic effects, except for benzodiazepines, which may reduce the antidepressant effects of ketamine. Despite these limitations, the outcomes from our analysis largely align with published literature, offer preliminary novel data relating to comorbid diagnoses of MDD and PTSD, and provide support for future research with larger trials to address any differences.

## 5. Conclusion

This study assessed the changes in biological and psychological parameters for MDD and comorbid MDD+PTSD participants in low-dose oral ketamine treatment trials. There is urgent need for clinical advancements of rapid, robust, and efficacious pharmacotherapeutics for mental health disorders. Low-dose oral ketamine offers great potential as a rapid-acting translational therapeutic treatment for MDD and MDD+PTSD. Our results suggest an etiological and biological overlap between MDD and PTSD and overlapping benefits for treatment of these disorders separately, and concurrently. Additionally, blood-based biomarkers have a place in understanding the treatment response to ketamine and further study with larger cohorts across longer time periods are needed. Overall, our study continues to support evidence that low-dose oral ketamine is efficacious and provides rapid, robust significant psychological improvements for challenging to treat disorders such as MDD and MDD+PTSD.

## Glossary

MDD: major depressive disorder
PTSD: post-traumatic stress disorder
BDNF: brain-derived neurotrophic factor
VEGF: vascular endothelial growth factor A
FUP1: follow-up one-week
DASS-21: Depression, Anxiety, and Stress Scale
SOFAS: Social and Occupational Functioning Assessment Scale
BSS: Beck Scale for Suicidal Ideation
WHO-5: World Health Organisation Well-Being Index
HAM-D: Hamilton Depression Rating Scale
MADRS: Montgomery-Asberg Depression Rating Scale
PCL-5: Post-traumatic Stress Disorder Checklist for DSM-5

## Data Availability

All data produced in the present study are available upon reasonable request to the authors.

## Acknowledgements

The study team would like to gratefully acknowledge and thank all the participants who volunteered their time to be part of this work and the Clinical Research Unit at the Thompson Institute, UniSC, for their support of the clinical programs underlying this study. Specifically, thank you to team members Megan Dutton, Monique Jones, Fiona Randall, Jim Lagopoulos, Adem T. Can, Cyrana C Gallay and Grace Forsyth.

## Author contribution: CRediT statements

AB: Conceptualization, Methodology, Formal analysis, Writing – original draft.

CD: Supervision, Writing – Review & Editing

DH: Funding Acquisition, Writing – Review & Editing

BQ: Conceptualization, Methodology, Formal analysis, Supervision, Writing – Review & Editing

## Ethical statement

All studies in this manuscript were performed in compliance with relevant laws, regulations, and institutional guidelines and received appropriate institutional committees approval before commencement (Metro North Health Human Ethics Committee (HREC/2019/QPCH/53437 and HREC/18/QPCH/288) and the UniSC Ethics Committee (A211537 and A181190, with trails prospectively registered with the Clinical Trials Registry of Australia and New Zealand under ACTRN12621000429853 and ACTRN12618001965291). Informed consent was obtained for research from all participants, including the use of their data and biological materials. Consent was given without coercion, and the privacy rights of participants was always observed.

## Declaration of generative AI

Generative AI was not used in any way for this paper.

---

**Supplementary Table 1:** Participant recruitment information, as well as inclusion and exclusion criteria for this study.

| Participant recruitment | Inclusion criteria | Exclusion criteria |
| --- | --- | --- |
| Study participants were recruited through research connections to local psychiatrists, psychologists, and general practitioners (GPs), through research staff visits to medical clinics and by telephone conversations. Approved advertisement material was supplied to GP clinics and local hospitals. Most participants were civilians. | A confirmed current diagnosis of MDD or PTSD from a qualified psychologist, utilising MADRS or HAM-D for depression diagnosis, and CAPS-5 for PTSD diagnosis. Ability to provide written informed consent, fluent in the English language. Must be able to tolerate ketamine treatment, rating scales and blood tests. Remain in the study and be monitored on an ongoing basis. Did not satisfy any of the exclusion criteria. | Psychosis, mania, acute suicidality or history of ketamine abuse, history of epilepsy or unexplained seizures, cardiovascular disease or recent myocardial infarct (within previous 6 months), history of stroke, hypertension (resting blood pressure >150/100), weight >150kg, history of cerebral trauma or traumatic brain injury, abnormal liver function test results, previous adverse reaction to ketamine, pregnancy, current breastfeeding or planning a pregnancy, and concurrent engagement in other clinical trial interventions. |

**Supplementary Table 2A.**
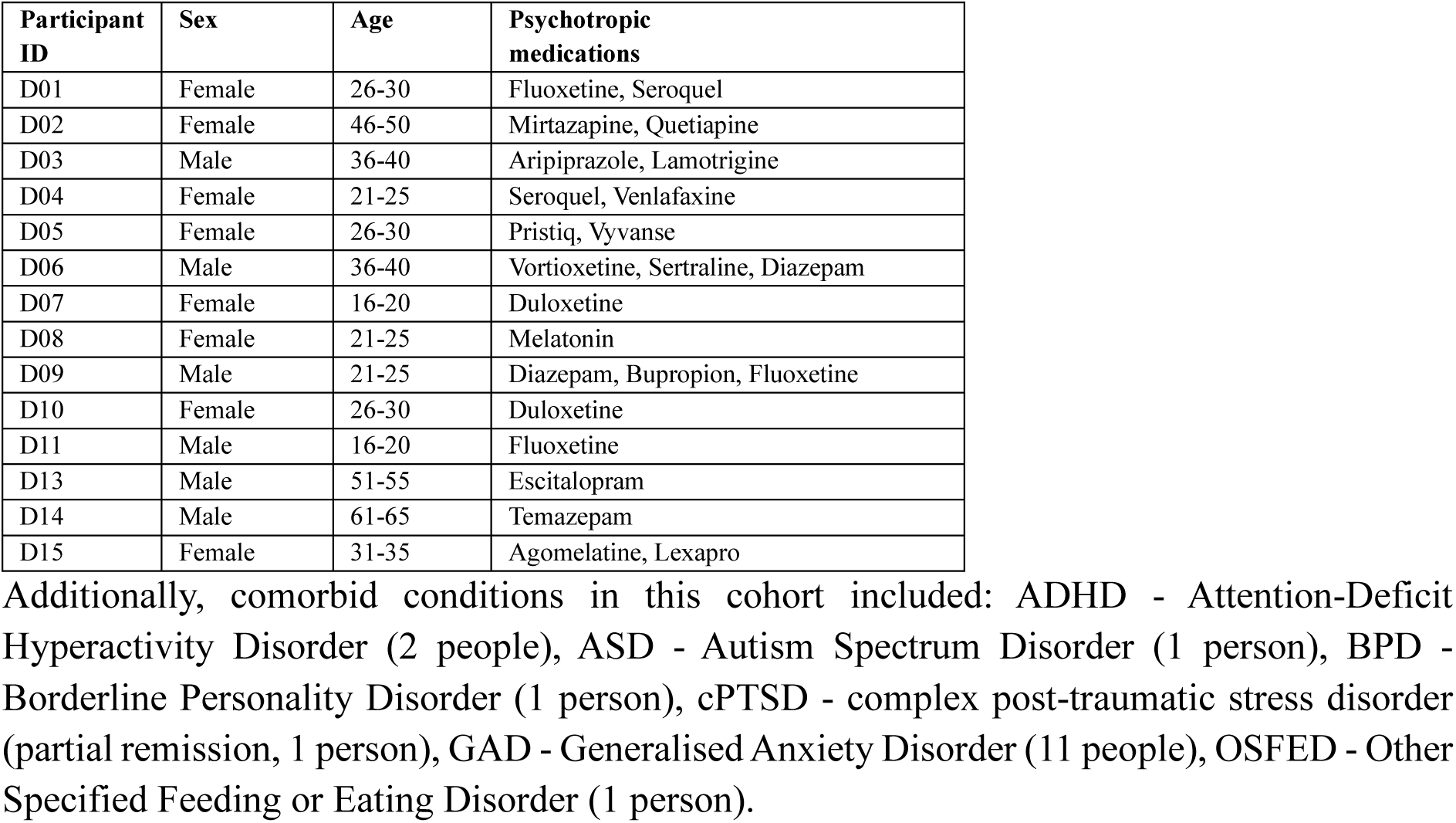
Demographic data for study participants (n=15). All participants met criteria for Major Depressive Disorder (MDD). (MDD participants).

| Participant ID | Sex | Age | Psychotropic medications |
| --- | --- | --- | --- |
| D01 | Female | 26-30 | Fluoxetine, Seroquel |
| D02 | Female | 46-50 | Mirtazapine, Quetiapine |
| D03 | Male | 36-40 | Aripiprazole, Lamotrigine |
| D04 | Female | 21-25 | Seroquel, Venlafaxine |
| D05 | Female | 26-30 | Pristiq, Vyvanse |
| D06 | Male | 36-40 | Vortioxetine, Sertraline, Diazepam |
| D07 | Female | 16-20 | Duloxetine |
| D08 | Female | 21-25 | Melatonin |
| D09 | Male | 21-25 | Diazepam, Bupropion, Fluoxetine |
| D10 | Female | 26-30 | Duloxetine |
| D11 | Male | 16-20 | Fluoxetine |
| D13 | Male | 51-55 | Escitalopram |
| D14 | Male | 61-65 | Temazepam |
| D15 | Female | 31-35 | Agomelatine, Lexapro |

Additionally, comorbid conditions in this cohort included: ADHD - Attention-Deficit Hyperactivity Disorder (2 people), ASD - Autism Spectrum Disorder (1 person), BPD - Borderline Personality Disorder (1 person), cPTSD - complex post-traumatic stress disorder (partial remission, 1 person), GAD - Generalised Anxiety Disorder (11 people), OSFED - Other Specified Feeding or Eating Disorder (1 person).

**Supplementary Table 2B.**
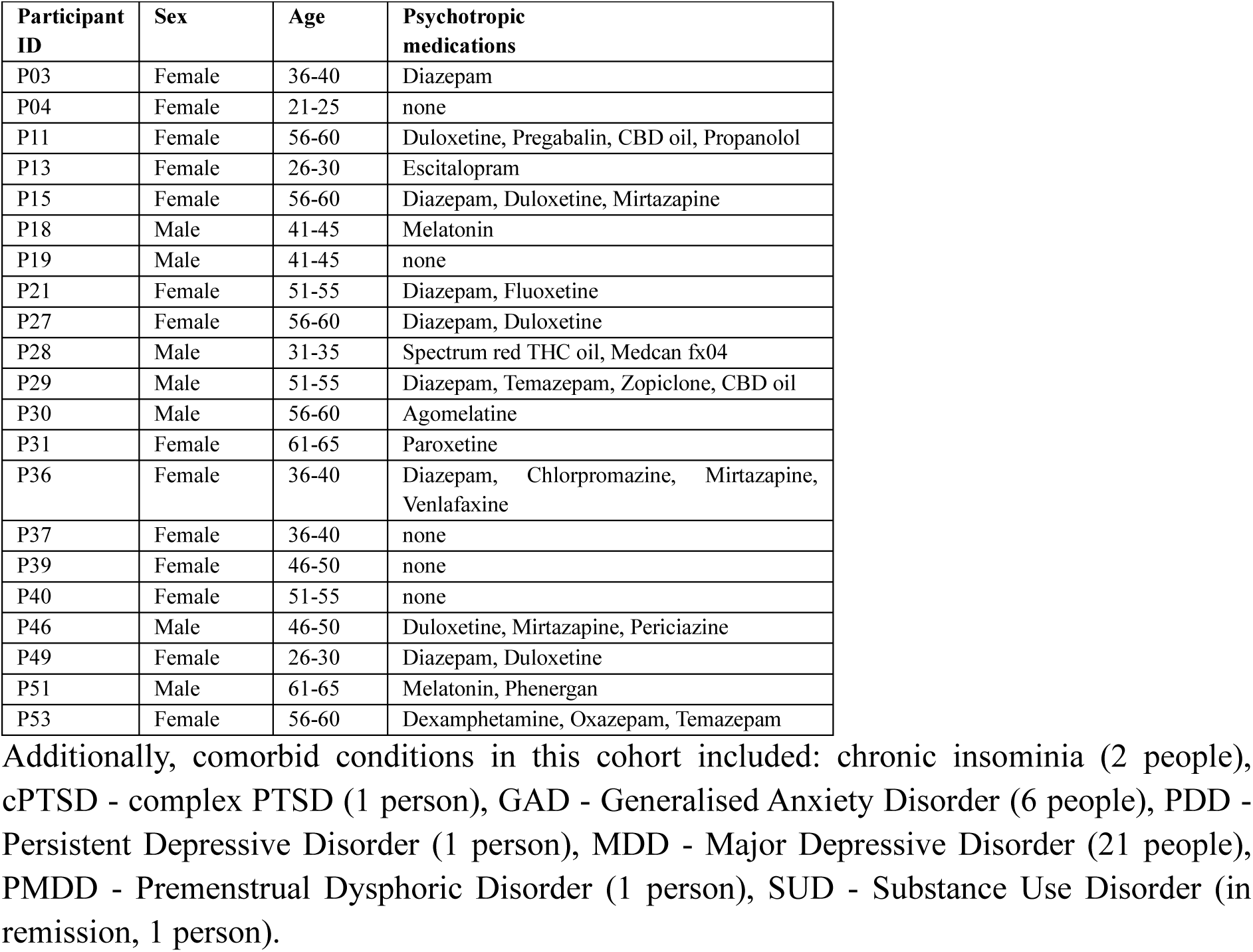
Baseline demographic data for the PTSD study participants (n=25). All participants met criteria for post-traumatic stress disorder (PTSD). (MDD+PTSD (n=21) participants).

